# Artemisinin Partial Resistance Mutations in Zanzibar and Tanzania Suggest Regional Spread and African Origins, 2023

**DOI:** 10.1101/2025.03.22.25323829

**Authors:** Sean V. Connelly, Julia G. Muller, Mohamed Ali, Billy E. Ngasala, Wahida Hassan, Bakari Mohamed, Kyaw L. Thwai, Jacob M. Sadler, Jacob Marglous, Abebe A. Fola, Abdallah Zacharia, Shija J. Shija, Safia Mohammed, Msolo C. Dominick, Hamza Said, Editruda E. Peter, Melic Odas, Isaack J. Rutha, Mwanaidi Nwange, Karamoko Naire, Shazia Ruybal-Pesántez, Robert Verity, Rebecca Crudale, Varun Goel, Barbara B. Choloi, Anders Björkman, Jeffrey A. Bailey, Jessica T. Lin, Jonathan J. Juliano

**Author notes:** Corresponding Author: Jonathan Juliano, MD, 110 Mason Farm Rd., CB#7036, University of North Carolina, Chapel Hill, NC 27599, United States of America. Co-first authors (Sean V. Connelly, Julia G. Muller). Co-senior authors (Jeffrey A. Bailey, Jessica T. Lin, Jonathan J. Juliano).

## Abstract

Artemisinin partial resistance (ART-R), driven by Plasmodium falciparum K13 mutations, threatens malaria control. Zanzibar is vulnerable to ART-R spread but lacks recent molecular surveillance. We sequenced samples in Zanzibar and mainland Tanzania collected in 2022-2024. K13 mutations (P441L, A675V) were found in 2/1440 Zanzibar participants and 6/3762 (R561H, P441L) in mainland Tanzania. K13 mutations appear to be of African origin and spreading regionally based on whole genome sequencing data. Frequent parasite importation appears to maintain partner drug mutation frequencies similar to the mainland, where artemether-lumefantrine selects for mutations favoring artesunate-amodiaquine sensitivity. Ongoing molecular surveillance remains essential to track these patterns.

**Summary:** K13 mutations linked to artemisinin resistance were uncommon in Zanzibar and mainland Tanzania in 2022–2024. Whole genome sequencing suggests an African origin with regional spread of mutations. Continued surveillance is critical to monitor evolving resistance and drug selection dynamics.

## Introduction

East Africa is facing a potential public health crisis with the emergence of artemisinin partial resistance (ART-R) in the region [1]. ART-R can be mediated by mutations in *Plasmodium falciparum* kelch 13 (K13) gene. These mutations have been correlated with *in vivo* phenotypes of ART-R, prolonged parasite clearance, and longer clearance half-lives. K13 polymorphisms are now found at a moderate prevalence in certain parts of Rwanda, Ethiopia, Tanzania, and Uganda, with isolates containing the mutations found sporadically across the entire East Africa region [1]. A concern is that ART-R will set the stage for partner drug resistance to emerge in Africa, as more parasites will be exposed to partner drug monotherapy as a part of artemisinin combination therapies (ACTs) [1]. Mutations that confer tolerance to partner drugs exist in Africa, such as polymorphisms in *P. falciparum* multi drug resistance protein 1 (MDR1) and chloroquine resistance transporter (CRT), which may also potentiate the spread of ART-R [2].

Zanzibar is a semi-autonomous archipelago off the coast of Tanzania, with an independent malaria control program and Ministry of Health. Unlike mainland Tanzania, where some regions have moderate to high levels of transmission, Zanzibar has been a pre-elimination setting for almost a decade, with low levels of transmission and susceptibility to outbreaks [3]. Much of the malaria in Zanzibar is thought to be imported from the adjacent Tanzanian mainland [3]. Zanzibar is likely at increased risk for ART-R given emergence of the validated R561H mutation in Kagera region in northwestern Tanzania [4], the relatively low immunity from years of malaria control (resulting in a greater proportion of infected individuals being symptomatic, seeking treatment and leading to increased drug pressure [5]), and the presence of competent Anopheline vectors.

Previous molecular surveillance in 2017 at 14 health centers throughout both islands in Zanzibar showed no K13 polymorphisms and low levels of mutations in genes associated with reduced susceptibility to partner drugs [6]. This study leverages comprehensive sampling of malaria cases across Unguja, Zanzibar, and selected regions of mainland Tanzania to assess the extent that emerging drug resistance polymorphisms in mainland Tanzania have spread eastward and threaten ACT efficacy in Zanzibar.

## Methods

We successfully sequenced 5,202 samples, collected from malaria cases diagnosed by rapid diagnostic tests, using molecular inversion probes (MIPs) targeting key antimalarial resistance mutations collected at 100 health facilities in Unguja, Zanzibar, and from 29 health facilities in 24 districts from mainland Tanzania between May 2022 and January 2024 (**Figure 1**). The mainland health facilities were selected in regions most commonly reported as travel destinations by Zanzabaris with malaria that were investigated the previous year by the Zanzibar Malaria Elimination Program (ZAMEP). Zanzibari health facilities were selected using a stratified random sampling approach to draw evenly from urban and rural areas and sample roughly half of malaria cases across the main island of Unguja, with the goal of enrolling every malaria case presenting at the participating clinic. Health facility staff were trained by the research team to consent participants, collect dried blood spots (DBS) on Whatman 3 filter paper, and conduct a brief survey of clinical, behavioral, demographic, and travel-related questions. Consents, case report forms, and DBS were packaged by the health facility staff and collected by the research team during site visits and audits. Packets were returned to the Muhimbili University of Health and Allied Sciences (MUHAS) laboratory in Bagamoyo, Tanzania where consents were confirmed, survey data were scanned into an electronic database using ABBYY Flexcapture software, and DBS punched into 96 deep well plates (3 punches per sample). All data scanned were manually checked and corrected as needed by research staff. The number of samples sequenced from each health facility is shown in **Table S1** and the seasonality of collections of sequenced samples in Zanzibar and the mainland are shown in **Figure S1**.

**Figure 1.**
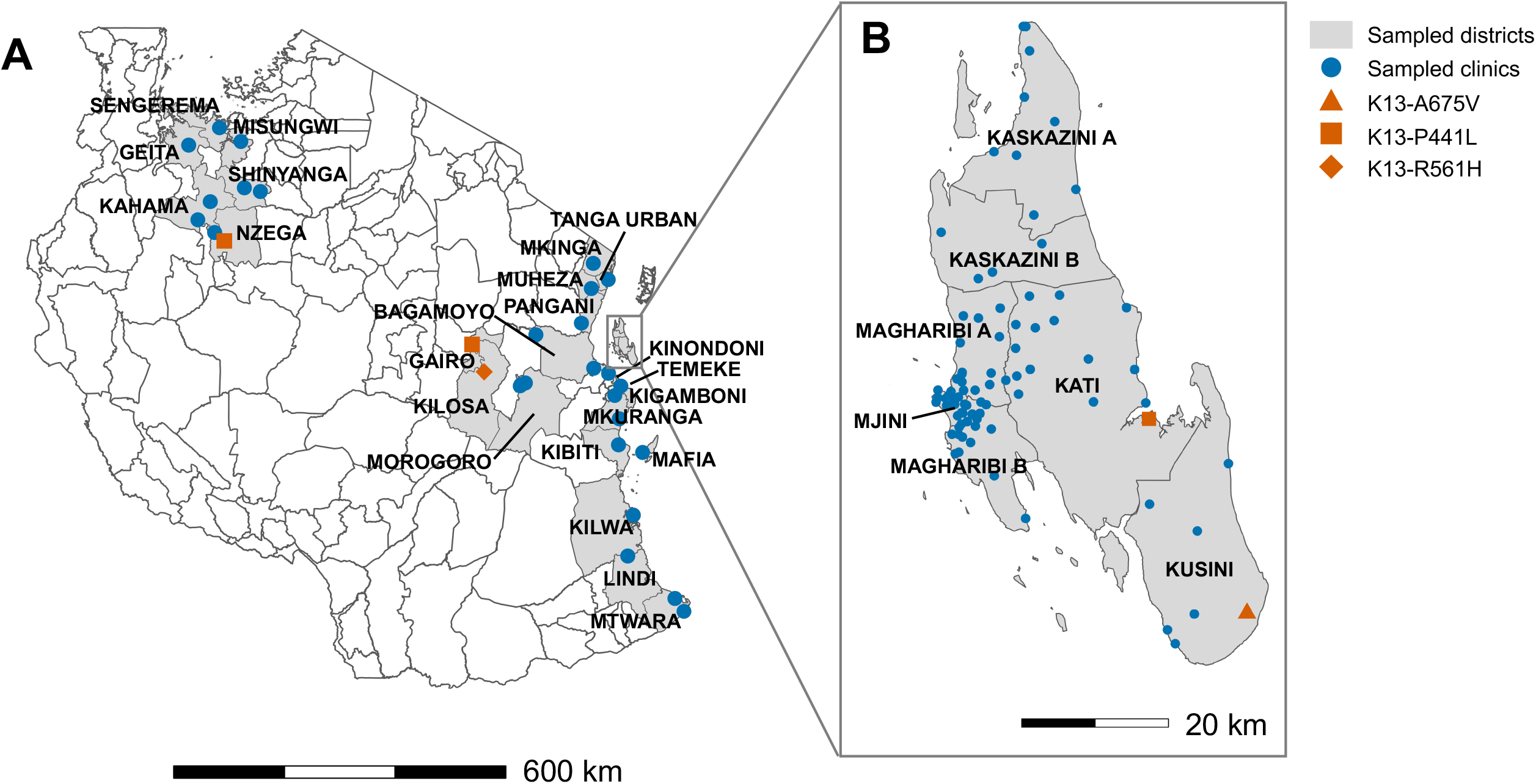
Map of health facilities for the study and locations of isolates with K13 artemisinin partial resistance mutations. Panel A shows locations of health facilities in the mainland and Panel B shows health facilities in Unguja, Zanzibar. In total, 3,762 samples from 29 clinics in mainland Tanzania and 1,440 from 100 clinics in Zanzibar were genotyped.

Samples were shipped to the University of North Carolina, where DNA was extracted from the DBS using a Chelex extraction method. Parasitemias were estimated using an ultrasensitive real-time PCR targeting the *varats* gene using mocked blood spots at various concentrations as controls [7]. The mock blood spots were generated by mixing culture grown 3D7 (MRA-102, BEI, Manasas, VA) with whole blood at specific parasitemias, which was spotted and dried on filter paper. Extracted DNA was then sent to Brown University to undergo MIP capture and sequencing of 64 targeted antimalarial resistance polymorphism loci in *P. falciparum* (see **Table S2** for the complete list of markers) [8]. Resulting data were analyzed using MIPtools software with freebayes variant calling (https://github.com/bailey-lab/MIPTools) [8]. Controls for each MIP capture and sequencing included DNA from 3D7 and 7G8 (MRA-102G and 152G, BEI) as well as no template and no probe controls. Variant calling was conducted as previously described [8]. Samples were filtered based on unique molecular identifier (UMI) coverage of greater than or equal to 5 UMIs per locus. Mutation prevalence was determined by dividing the sum of the mixed genotype and pure resistant individuals by the total amount of pure resistant, mixed and reference individuals. Prevalence was stratified by region, and by travel history status among cases from Zanzibar (mainland Tanzanian samples, Zanzibari travelers or Zanzibari non-travelers). A 95% confidence interval was calculated with the Pearson-Klopper exact method. Mutant K13 isolates underwent whole genome sequencing (WGS), quality control and were compared with publicly available WGS data from Africa and Southeast Asia, as described in the **Supplementary Materials**.

## Results

Demographics of participants are shown in **Table S3**. Participants exhibited some demographic differences between regions; namely, the majority of malaria cases in Zanzibar were in males (67.0% in Zanzibar vs. 46.7% in mainland), and malaria cases in mainland Tanzania tended to be younger-aged children. Mainland Tanzania malaria cases also exhibited higher median parasite density compared to those from Zanzibar (6,660 p/µL in mainland vs. 3,810 p/µL in Zanzibar). Among the Zanzibari participants sequenced, 569/1440 (39.5%) reported travel to the mainland in the preceding 28 days.

A total of 1,440 Zanzibari and 3,762 mainland samples were successfully genotyped. Prevalences of all K13 polymorphisms detected in this study are in **Table S4**, and WHO validated or candidate ART-R K13 polymorphisms are highlighted in **Figure 1**. Among the Zanzibari isolates, we identified 2 samples with WHO validated or candidate ART-R K13 polymorphisms. One sample collected in Kati district (central Unguja) from a female who did not identify as a permanent Zanzibari resident and had recently traveled to Kigoma district (western Tanzania, bordering the Democratic Republic of the Congo) contained the P441**L** mutation. The second was collected in Kusini (southeast Unguja) from a student who had not traveled in the last 28 days and contained the A675**V** mutation. Interestingly, R561**H**, which has been detected throughout the Great Lakes region, including in Tanzania, was not observed in Zanzibar. On the mainland, 3 isolates contained the P441**L** mutation (2 from Gairo in the central Morogoro region, 1 from the Nzega district of Tabora in northwest Tanzania) and 3 contained the R561**H** mutation, all from the Kilosa district of Morogoro. The prevalence of molecular markers for all other antimalarials was similar between Zanzibar and the mainland (**Figure S2** and **Table S5**), including commonly identified wild-type MDR1 **N**86Y allele, associated with increased tolerance to lumefantrine, and uncommon CRT K76**T** and MDR1 N86**Y** mutant alleles associated with tolerance to amodiaquine (AQ). However, there were some subtle differences between our previous report leveraging samples from 2016-2018 and current estimated prevalences from samples collected 2022-2024 [9]. For example, dihydropteroate synthase (DHPS) A581**G** mutation increased in prevalence in Zanzibar from 0.04 (8/181, 95% CI: 0.02–0.09) to 0.12 (142/1181, 95% CI: 0.10-0.14) (p = 0.004).

We assessed the extended haplotypes around K13 from all mutant isolates detected in the study by conducting whole genome sequencing, comparing to publicly available African and Asian genomes containing mutant R561**H**, A675**V**, and P441**L** (**Table S6**). We generated high-quality genomes for eight (3 R561**H**, 1 A675**V**, and 4 P441**L**) isolates. The R561H isolates from the Morogoro region cluster with the previously described TZ2 haplotype identified in Kagera in 2021 (**Figure S3**) [4]. The Zanzibari isolate with A675**V** clustered with both mutant and wild-type isolates from Africa, and not with A675**V** mutant isolates from Asia (**Figure S4**). Lastly, for P441**L**, the 3 isolates from mainland Tanzania and single isolate from Zanzibar cluster together and are more closely related to African isolates, rather than Asian P441**L** mutant isolates (no mutant African P441**L** genomes were available) (**Figure 2**). These preliminary findings suggest regional transmission of R561**H** and P441**L** clones is occurring within Tanzania, including to Zanzibar. These are the first A675**V** and P441**L** isolates sequenced from Africa, and data support novel African haplotypes rather than haplotypes from Asia. One of the two cases of K13 mutations in Zanzibar did have recent travel that suggests importation from Kigoma, but additional sequencing of isolates outside of Tanzania to assess extended haplotypes around the K13 locus is needed to better understand if these parasites are related to parasites carrying the same mutation in other regions of East Africa.

**Figure 2.**
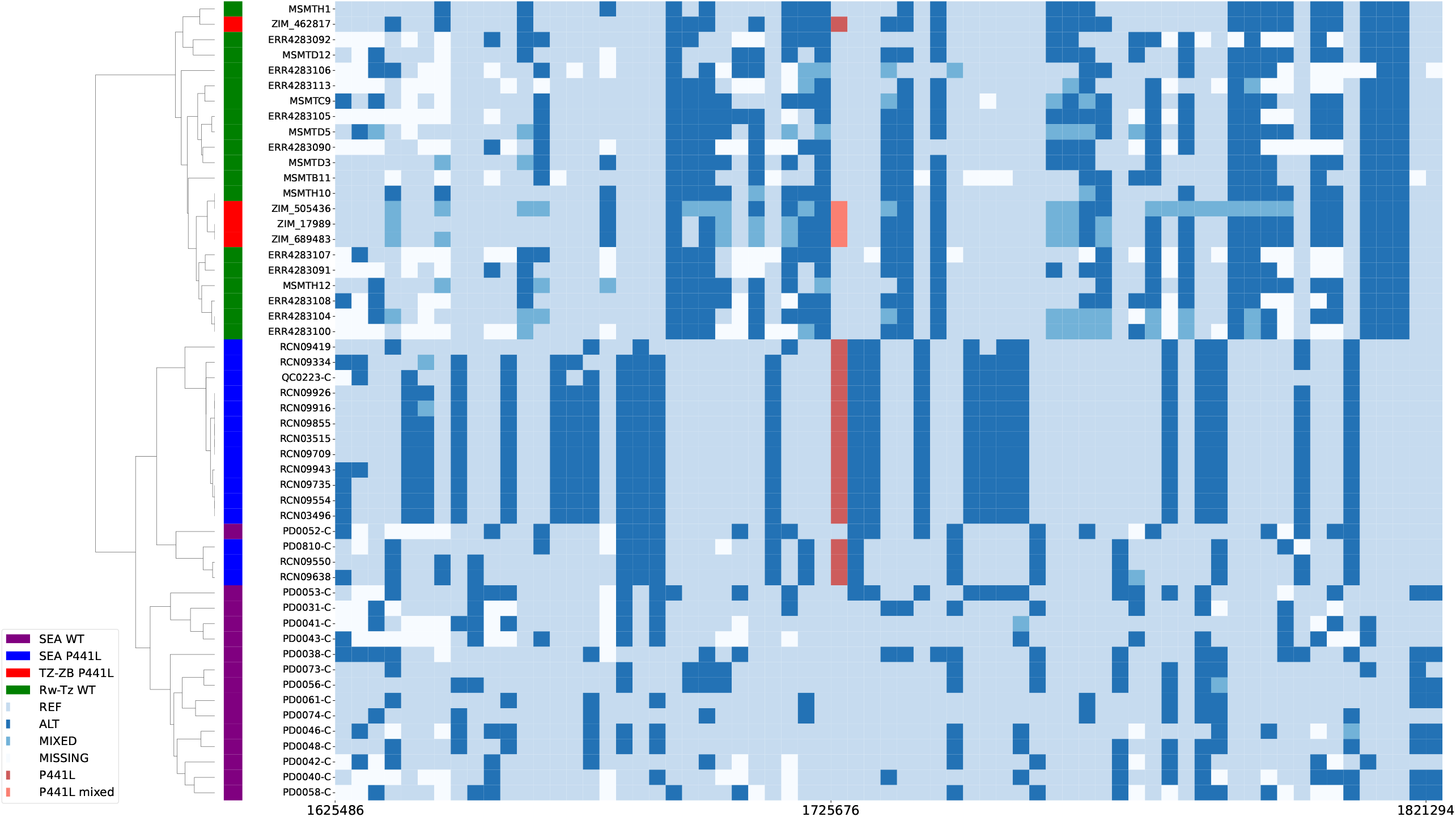
Extended Haplotype Analysis of P441L K13 Mutant Isolates from Zanzibar and Tanzania. Extended haplotypes 50kb upstream and downstream of the P441L mutation (chr13:1725676) were compared between mainland Tanzania, Zanzibari, and publicly available Southeast Asian (SEA) genomes. Mutants from the mainland Tanzania and Zanzibar (red) cluster with wildtype parasites from mainland Tanzania (green: MSMT-) and appear distinct from mutant P441L samples from Asia (blue: PD-, Thailand; QC-/RCN-, Myanmar). The parasites from this study also appear distinct from wildtype SEA (purple: PD-Thailand) and Rwandan samples (green: ERR-). Taken together, this suggests transmission between mainland Tanzania and Zanzibar, and an African origin of these P441L mutants.

## Discussion

ART-R mutations are now being seen in Zanzibar and raise concern over the spread of artemisinin partial resistance to this receptive low transmission region. Similar to previous work, ART-R polymorphisms remain rare and heterogeneously distributed across the mainland sites, but are detectable [4]. This study did not sample in the Kagera region where K13 mutations have been commonly found [4,10]. The A675**V** mutation was originally described in Uganda, but has spread regionally and has been reported in Tanzania previously [4]. The P441**L** mutation has also already been described in multiple locations in East Africa [4,11], and now Southern Africa. However, this is the first reported detection of this mutation in Tanzania.

Zanzibar continues to use artesunate-amodiaquine (AS-AQ) as its first-line antimalarial, while mainland Tanzania uses artemether-lumefantrine (AL). AQ and lumefantrine have differential selection on key antimalarial resistance polymorphisms, in particular MDR N86Y and CRT K76T, where lumefantrine selects for the wild type and AQ selects for the mutant. The finding of a resistance pattern more similar to regions that use AL (such as the mainland, **Figure S2**) on Zanzibar strongly suggests that importation is a major driver shaping the parasite population, as the opposite signatures of selection on antimalarial resistance alleles would be expected based on first-line therapy in Zanzibar, as seen with AS-AQ use in Uganda.

Given the large numbers of clinics, the number of samples sequenced to date does not enable a refined understanding of differences on a local scale. Continued sequencing of isolates is ongoing. We are also limited by the relatively few number of sites from the mainland where samples were collected. This could be overcome by accessing samples from other large studies, such as school-aged children surveys [12] or other major surveillance efforts. Currently, we cannot deduce the source of these K13 mutations. Eventually, as more whole genomes become available from African isolates with different mutations, it will be possible to use analysis of K13 flanking haplotype to determine if these are imported cases from other areas of Tanzania and Africa, or if they are independent origins arising locally [4].

The emergence of ART-R is concerning for the longevity of ACTs in Africa. To date, efficacy generally remains high despite the presence of K13 mutations [10,13]. However, continued importation and further spread of these resistance mutations could threaten ACT efficacy in Zanzibar. To date, there is no evidence of high-level resistance to lumefantrine or amodiaquine, the two most common partner drugs used in Africa, but isolated reports of clinical failure and trends toward higher rates of reduced sensitivity to lumefantrine are concerning [14,15]. In Zanzibar, we identified two isolates with K13 mutations. It is possible that the use of AS-AQ in Zanzibar has helped to reduce onward transmission of K13 mutations introduced, given the favorable partner drug tolerance profile. For now, widespread routine molecular surveillance for polymorphisms associated with antimalarial resistance remains the best early warning sign for potential clinical problems with ACTs in the future.

## Acknowledgements

We thank the study participants who donated their time and the health facility staff who collected samples for the study. We thank the ZAMEP district malaria surveillance officers who were instrumental to the successful implementation of the study. The following reagent was obtained through BEI Resources, NIAID, NIH: *P. falciparum*, Strain 3D7, MRA-102, contributed by Daniel J. Carucci. The following reagent was obtained through BEI Resources, NIAID, NIH: Genomic DNA from *P*.*falciparum*, Strain 3D7, MRA-102G, contributed by Daniel J. Carucci. The following reagent was obtained through BEI Resources, NIAID, NIH: Genomic DNA from *P. falciparum*, Strain 7G8, MRA-152G, contributed by David Walliker.

## Ethics Statement

The study was approved by the institutional review board (IRB) in Zanzibar (ZAHREC/01/EXT/July/2023/03), at MUHAS (MUHAS-REC-02-2024-1168), and at UNC (21-1966). Written informed consent, parental consent and/or assent were provided by all participants.

Sequencing data are available at PRJNA1271114. Metadata are available at https://doi.org/10.15139/S3/EU9GGW. Custom scripts are available at https://github.com/sconnelly007/ZIM_ART_R.

## Author contributions

Conception and design: JJJ, JTL, BEN, JAB, MA, WH, AB; Data acquisition, analysis and interpretation: SVC, JGM, MA, WH, BM, JMS, JM, AF, KN, AZ, MCD, RC, KLT, VG, BBC, EEP, MO, IJR, MN, SRP, BV, HS; Drafting manuscript: SVC, JGM, AF, JTL, JJJ, JAB; Revision and final approval: All authors; Accountability: SVC, JGM, JAB, BEN, JTL, JJJ

