## Supplemental Material for "Artemisinin Partial Resistance Mutations in Zanzibar and Tanzania Suggest Regional Spread and African Origins, 2023"

**Whole Genome Sequencing and Haplotype Analysis**

*Library Preparation*: Genomic DNA from dried blood spots underwent 2 specific whole genome amplifications (sWGA) as previously described [1]. The replicate sWGAs were mixed, and whole genome sequencing libraries were prepared using the Watchmaker DNA Library Kit with Fragmentation (Watchmaker Genomics Inc., Boulder, CO). Libraries were pooled and sequenced using Illumina 2X150bp chemistry at the University of North Carolina High Throughput Sequencing Facility.

*Public Whole Genome Sequencing Data*: For analysis of extended haplotypes, public samples from Southeast Asia were downloaded from the Pf7 database [2], and samples from Rwanda and Tanzania were downloaded via SRA [3–5], as aligned read (bam) files. The Tanzania WGS data are derived from two studies: 1) a single-arm therapeutic efficacy study conducted between April 29 and September 1, 2022, at the Bukangara dispensary in the Karagwe district, located in the Kagera region of Tanzania [4], and 2) samples collected during the 2021 MSMT survey of the Kagera region [5]. Both study sites are within 100 km of Rukara, Rwanda, where ART-R was confirmed in 2018. All genome IDs are available in **Table S6**.

*Whole Genome Sequencing Analysis*: Following whole-genome sequencing, sequence reads were aligned to the Pf3D7 (version 3) reference genome with BWA-MEM. Both public and novel samples underwent quality control and variant calling using an optimized GATK4 pipeline for *P. falciparum* [6]. The pipeline is available at https://github.com/Karaniare/Optimized_GATK4_pipeline. To ensure high-confidence variant identification, SNPs and indels underwent filtering through variant quality score recalibration. The resulting multi-sample VCF was filtered via VCFtools: by sample to keep only samples with calls at more than 50% of sites, and by site to keep only biallelic SNP sites with calls for at least 90% of samples, and minimum Phred score of 30 [7]. We detected 3 heterozygous samples for P441L, 1 homozygous sample for P441L, 3 homozygous samples for R561H, and 1 homozygous sample for A675V.

*Genome Extended Haplotype Analysis*:
VCFs were converted to a genotype matrix via PySam, a Python wrapper for Samtools [8]. Further filtering was performed in Python to remove alternate alleles present at less than 10% within-sample allele frequency (WSAF). For visualization, Pf7 samples were filtered to only monoclonal samples by Fws of at least 0.95 [9]. Then, for each *k13* mutant (P441L, R561H, and A675V), haplotypes of mutant samples sequenced in this study were compared against the available Rwandan and Tanzanian R561H mutants and wild-type samples, as well as a random set of Pf7 wild-type (15) and mutant (15) isolates. Samples were clustered using the furthest point algorithm on a matrix of Manhattan distances, as implemented in Scipy [10], and visualized using Seaborn [11].

**Figure S1. Seasonality of samples genotyped on the mainland and Zanzibar**

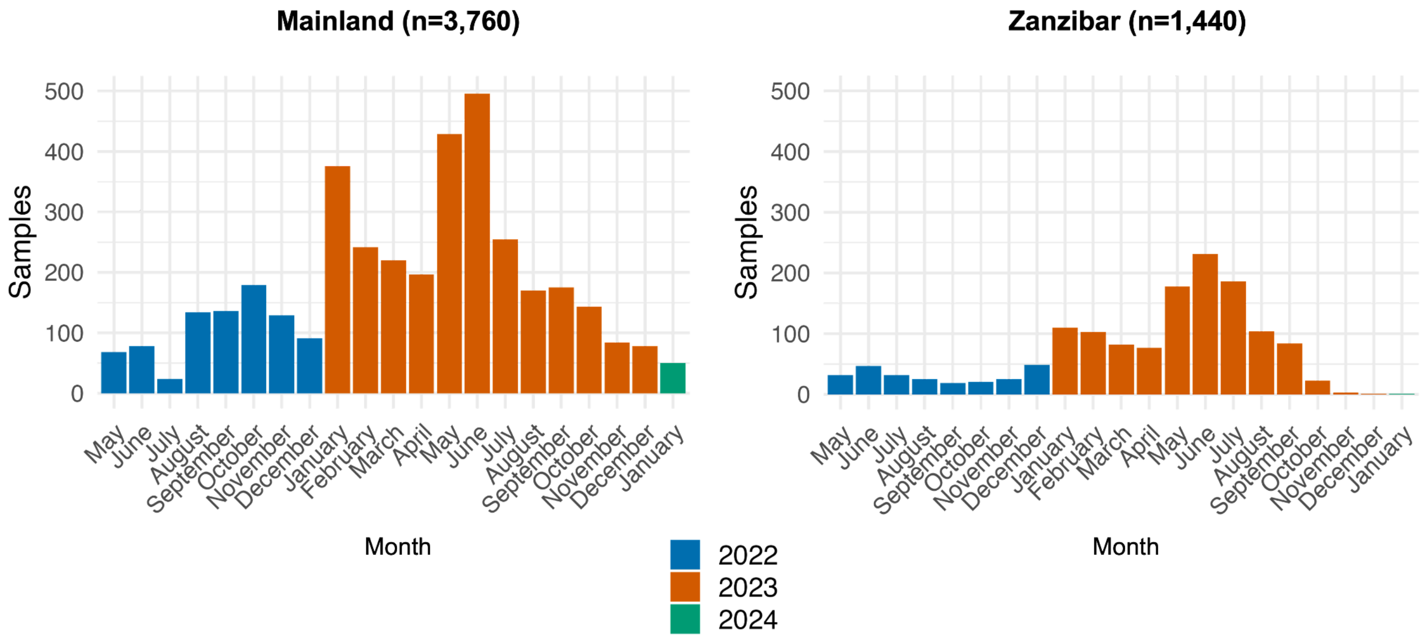

**Figure S2. Prevalence estimates for non-K13 antimalarial molecular markers in Zanzibar and mainland Tanzania.**

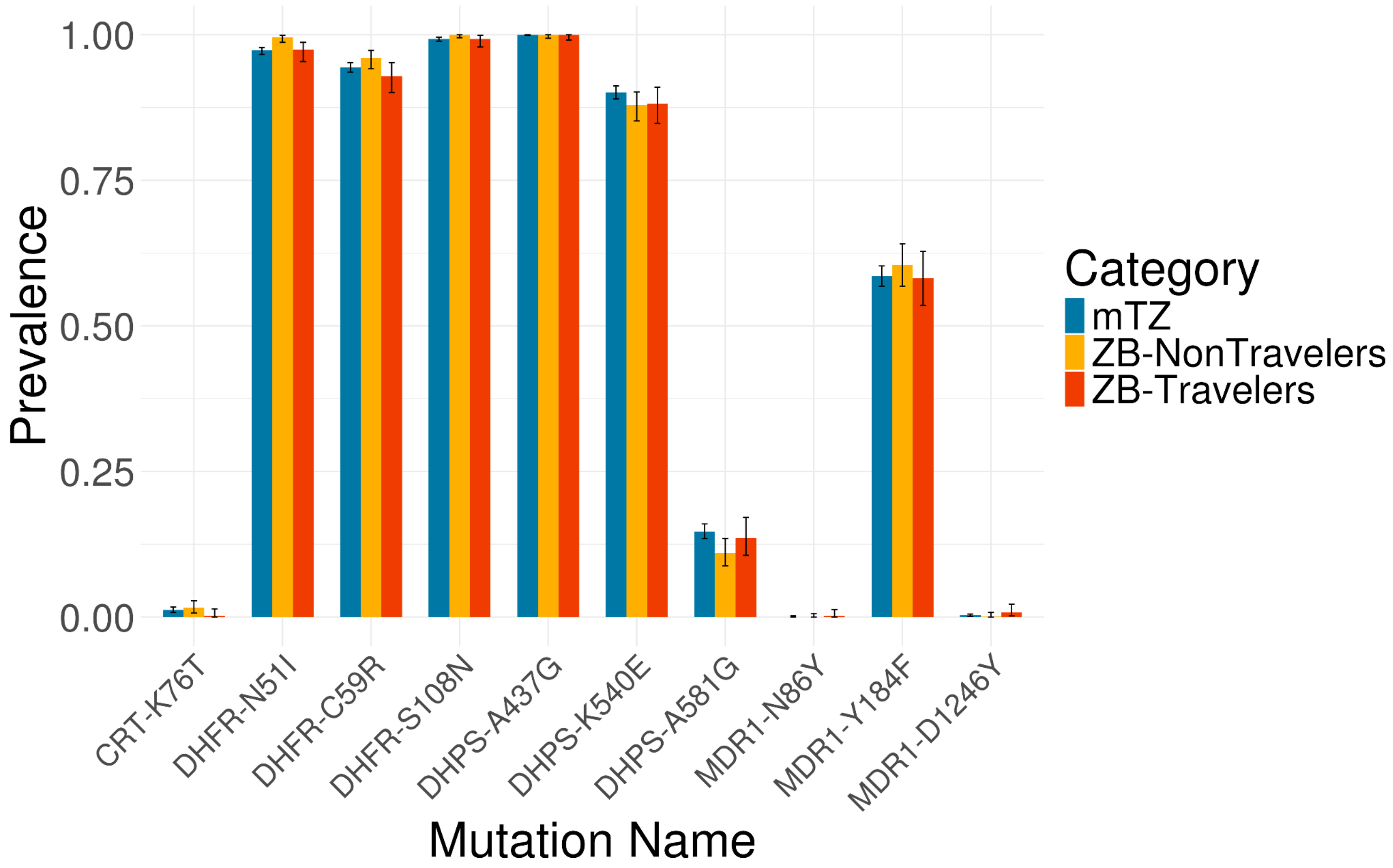

**Figure S3. Extended Haplotype Analysis of R561H K13 Mutant Isolates**. Extended haplotypes 100kb upstream and downstream of the R561H mutation (chr13:1725316) were compared between mainland (3) and publicly available genomes. The three mutant R561H mutant isolates (red) cluster with other sequenced mutant isolates from Africa, in particular TZ2 haplotypes (yellow) from Kagera, Tanzania, rather than Asian mutant (blue) or wild-type (purple) isolates from public data (PD-, Thailand; RCN-, Myanmar), or African TZ1 haplotypes (orange). African samples were collected in Tanzania (MSMT-)[5] and Rwanda (ERR-) [3]**.**

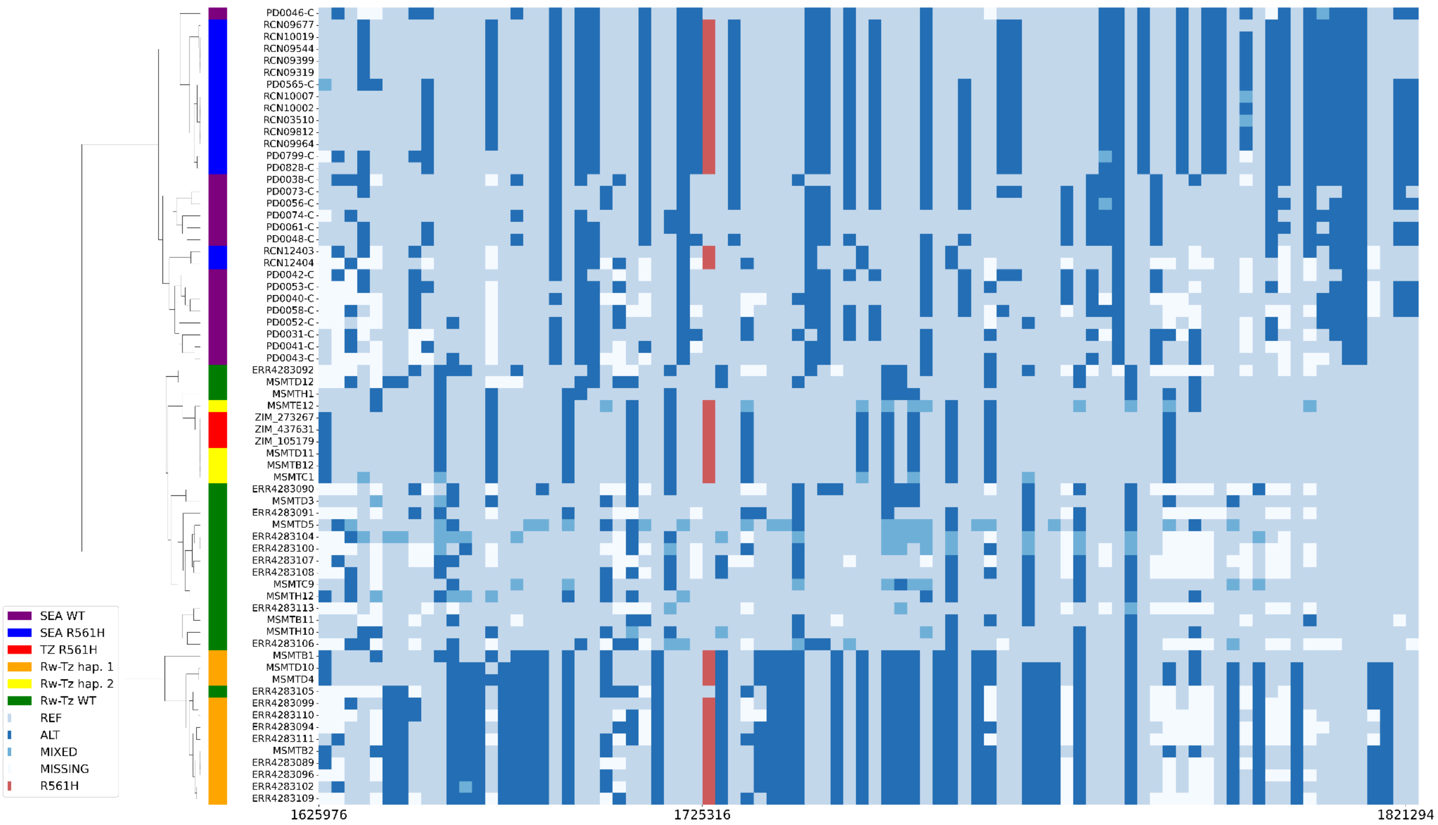

**Figure S4. Extended Haplotype Analysis of A675V K13 Mutant Isolates.** Extended haplotypes 100kb upstream and downstream of the A675V mutation (chr13:1724974) were compared between one Zanzabari isolate (red) and publicly available genomes. The Zanzibari isolate clusters with both A675V mutant African isolates (orange: a traveller from Uganda and a local parasite from Uganda [12,13]) and wild-type African isolates (green: MSMT- from Kagara, Tanzania, ERR- from Rwanda), rather than mutant A675V from Asia (blue: PD-, Thailand; QC-, Myanmar) from public data, suggesting an African origin.

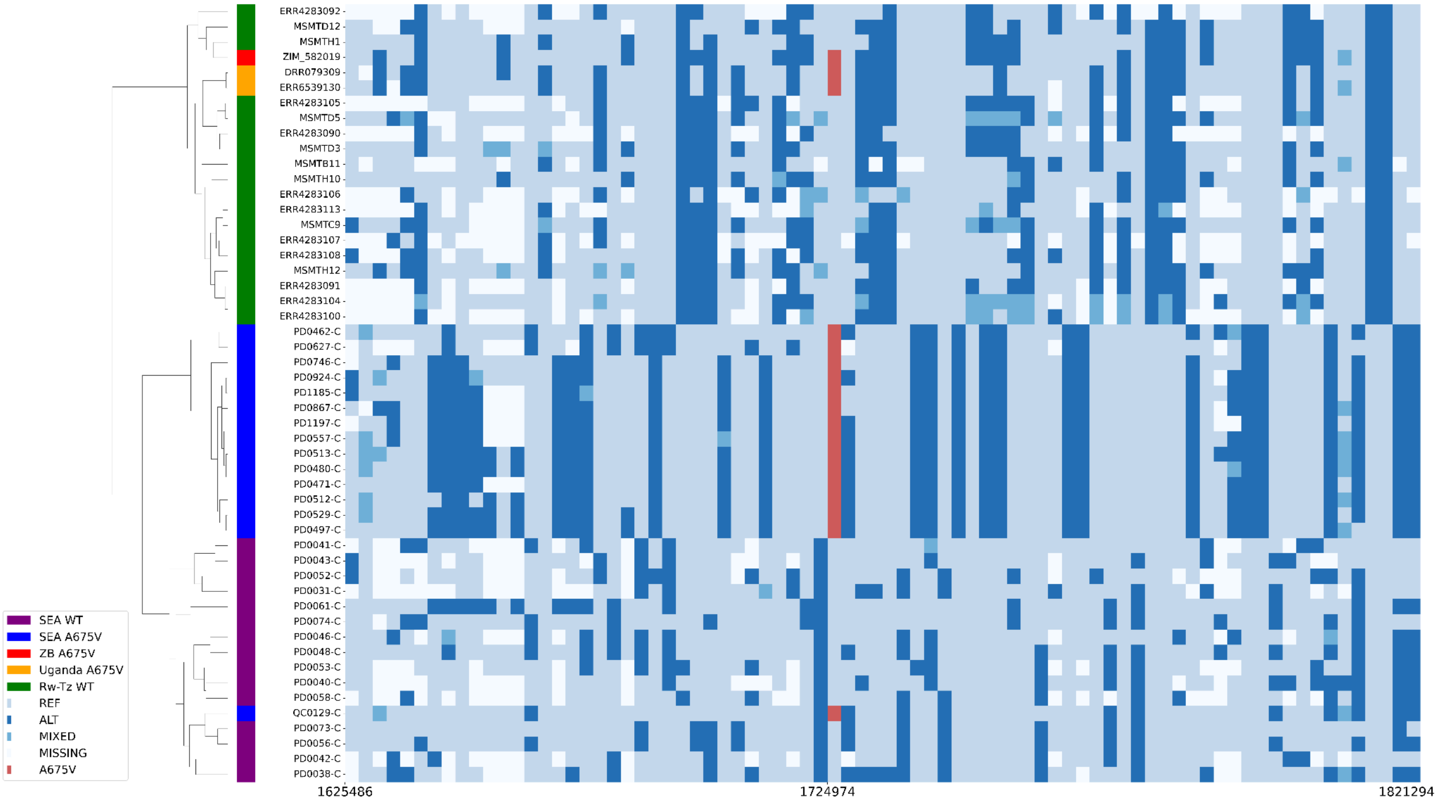

**Table S1. Number of samples genotyped at each clinic**

| Clinic | Clinic Name | District | Sample Size |
|---|---|---|---|
| TZ_C011 | Mkuzi Health Center | Muheza | 301 |
| TZ_C026 | Kibindu Health Center | Bagamoyo | 274 |
| TZ_C008 | Mikanjuni Health Center | Tanga | 255 |
| TZ_C003 | Mbande Dispensary | Temeke | 243 |
| TZ_C024 | Misungwi District Hospital | Misungwi | 220 |
| TZ_C006 | Mafia District Hospital | Mafia | 219 |
| TZ_C023 | Ushetu District Hospital | Ushetu | 203 |
| TZ_C004 | Kisiju Health Center | Mkuranga | 202 |
| TZ_C016 | Mnazi Mmoja Health Center | Lindi | 199 |
| TZ_C009 | Mkalamo Health Center | Pangani | 158 |
| TZ_C013 | Uhuru Health Center | Morogoro | 155 |
| TZ_C030 | Nkindu Dispensary (Nzega District) | Nzega | 135 |
| TZ_C021 | Shinyanga District Hospital | Shinyanga | 127 |
| ZB_C083 | Ziwani polisi | Mjini | 125 |
| TZ_C029 | Semembela Dispensary (Nzega District) | Nzega | 114 |
| TZ_C019 | Kilambo Health Center | Mtwara | 112 |
| TZ_C022 | Kahama District Hospital | Kahama | 112 |
| TZ_C001 | Kigamboni Health Center | Kigamboni | 89 |
| TZ_C015 | Kingolwira Health Center | Morogoro | 82 |
| ZB_C005 | Jangombe Mpendae | Mjini | 82 |
| TZ_C018 | Likombe Health Center | Mtwara | 80 |
| TZ_C028 | Ilekanilo Dispensary (Sengerema District) | Sengerema | 80 |
| ZB_C003 | Mafunzo | Mjini | 77 |
| TZ_C020 | Kambarage Health Center | Shinyanga | 73 |
| ZB_C006 | Raha Leo | Mjini | 65 |
| TZ_C027 | Unone Dispensary (Kilosa District) | Kilosa | 60 |
| TZ_C025 | Nyankumbu Health Center | Geita | 58 |

| | | | |
|---|---|---|---|
| TZ_C017 | Masoko | Kilwa | 52 |
| ZB_C009 | Utapoa Dispensary | Mjini | 51 |
| TZ_C002 | Bunju Health Center | Kinondoni | 47 |
| ZB_C001 | Jumbi Dispensary | Kati | 44 |
| ZB_C017 | Kizimkazi Mkunguni | Kusini | 43 |
| ZB_C050 | NOUR ALAA NOUR | Kaskazini A | 39 |
| TZ_C007 | Nyamisati Dispensary | Kibiti | 37 |
| ZB_C013 | Makunduchi | Kusini | 37 |
| ZB_C039 | Kizimkazi Dimbani | Kusini | 37 |
| ZB_C015 | Magogoni | Magharibi B | 36 |
| ZB_C082 | Kidutani | Mjini | 36 |
| ZB_C084 | New Asaakheri | Mjini | 34 |
| TZ_C012 | Gairo Health Center | Gairo | 33 |
| ZB_C002 | Chukwani | Kati | 32 |
| ZB_C004 | Bumbwisudi | Magharibi A | 32 |
| ZB_C085 | Shaurimoyo | Mjini | 30 |
| ZB_C011 | Mwera | Kati | 28 |
| ZB_C020 | Kisauni | Magharibi B | 24 |
| ZB_C018 | Minna Dispensary | Magharibi A | 23 |
| ZB_C012 | Kidongo Chekundu | Mjini | 22 |
| ZB_C016 | Kiembe Samaki | Magharibi B | 22 |
| ZB_C053 | TRIPLE J MEDICAL SERVICES | Magharibi B | 22 |
| ZB_C007 | Kianga Phcu | Magharibi A | 20 |
| ZB_C071 | Kivunge | Kaskazini A | 20 |
| ZB_C046 | CHARAWE | Kati | 19 |
| ZB_C008 | Al Tabib Dispensary | Mjini | 18 |
| ZB_C061 | Cheju | Kati | 18 |
| TZ_C010 | Mwanyumba Dispensary | Mkinga | 17 |
| ZB_C023 | Kidimni Phcu | Kati | 17 |
| TZ_C014 | Kihonda Health Center | Morogoro | 16 |

| | | | |
|---|---|---|---|
| ZB_C031 | Magirisi | Magharibi B | 16 |
| ZB_C096 | Care Clinic | Magharibi B | 16 |
| ZB_C038 | Miwani | Kati | 14 |
| ZB_C055 | KUNDI DISPENSARY | Mjini | 14 |
| ZB_C060 | MWEMBELADU HOSPITAL | Mjini | 14 |
| ZB_C067 | Uzini | Kati | 14 |
| ZB_C056 | SHASH DISPENSARY | Magharibi B | 13 |
| ZB_C099 | Sahal Hospital | Magharibi B | 13 |
| ZB_C033 | Mtofaani Phcu | Magharibi A | 12 |
| ZB_C066 | Jendele | Kati | 12 |
| ZB_C019 | Kitope Dispensary | Kaskazini B | 11 |
| TZ_C005 | Matimbwa Health Center | Bagamoyo | 9 |
| ZB_C027 | Pwani Mchangani Phcu | Kaskazini A | 9 |
| ZB_C029 | Upenja | Kaskazini B | 9 |
| ZB_C048 | CHWAKA | Kati | 9 |
| ZB_C058 | Habiba Dispensary | Magharibi A | 9 |
| ZB_C075 | Al-Shifaa Kazole Dispenary | Kaskazini B | 9 |
| ZB_C025 | Kauthar Specialized Clinic | Kaskazini A | 8 |
| ZB_C030 | Altwayybib Dispensary | Kaskazini B | 8 |
| ZB_C059 | BANDARINI | Mjini | 8 |
| ZB_C074 | Nungwi Phcu+ | Kaskazini A | 8 |
| ZB_C089 | Mbuzini Clinic And Dispensary | Magharibi A | 8 |
| ZB_C092 | Znz Clinic | Magharibi A | 8 |
| ZB_C010 | Matrekta | Magharibi B | 7 |
| ZB_C032 | Bwefum | Magharibi B | 7 |
| ZB_C037 | Afaa Medical Clinic | Magharibi B | 7 |
| ZB_C044 | AL-MANNA DISPENSARY | Kaskazini A | 7 |
| ZB_C047 | BWEJUU | Kusini | 7 |
| ZB_C026 | Mkokotoni Phcu | Kaskazini A | 6 |
| ZB_C034 | Muyuni | Kusini | 6 |

| | | | |
|---|---|---|---|
| ZB_C049 | BUMBWINI MISUFINI | Kaskazini B | 6 |
| ZB_C052 | AZHAR | Magharibi A | 6 |
| ZB_C080 | Hassan Clinic | Mjini | 6 |
| ZB_C035 | Mwera Pongwe | Kati | 5 |
| ZB_C040 | Bububu Dispensary | Magharibi A | 5 |
| ZB_C041 | Kendwa Phcu | Kaskazini A | 5 |
| ZB_C068 | Pongwe | Kati | 5 |
| ZB_C081 | J And M Dispensary | Mjini | 5 |
| ZB_C014 | Kitogani | Kusini | 4 |
| ZB_C021 | Jendele | Kati | 4 |
| ZB_C043 | MCHANGANI | Kati | 4 |
| ZB_C057 | ICB DISPENSARY | Mjini | 4 |
| ZB_C069 | Umbuji | Kati | 4 |
| ZB_C097 | Marie Stop Hospital | Magharibi B | 4 |
| ZB_C024 | Ndijani Mseweni | Kati | 3 |
| ZB_C051 | MNAZI MMOJA | Mjini | 3 |
| ZB_C062 | Gana | Kati | 3 |
| ZB_C076 | Fujoni | Kaskazini B | 3 |
| ZB_C088 | Kizimbani Phcu | Magharibi A | 3 |
| ZB_C022 | Koani Dispensary | Kati | 2 |
| ZB_C042 | MACHUI | Kati | 2 |
| ZB_C063 | Kiboje | Kati | 2 |
| ZB_C070 | Ahsana Dispensary | Kaskazini A | 2 |
| ZB_C079 | Michamvi | Kusini | 2 |
| ZB_C093 | Al-Hijri | Magharibi B | 2 |
| ZB_C028 | Dr. Metha Nungwi | Kaskazini A | 1 |
| ZB_C073 | Kijini Phcu | Kaskazini A | 1 |
| ZB_C087 | Al-Najid | Magharibi A | 1 |
| ZB_C100 | St. Camiliusn Dipensary | Magharibi B | 1 |

**Table S2. Targeted drug resistance markers and their polymorphisms utilizing Molecular Inversion Probes**

| Chromosome | Position | Gene | AA Position | Amino Acid Change | Mutation | Ref_Resistant? | Gene ID |
|---|---|---|---|---|---|---|---|
| chr14 | 2,481,070 | arps10 | V127 | Val127Met | arps10-V127M | No | PF3D7_1460900.1 |
| chr1 | 267,306 | atp6 | A623 | Ala623Glu | atp6-A623E | No | PF3D7_0106300 |
| chr1 | 268,386 | atp6 | L263 | Leu263Glu | atp6-L263E | No | PF3D7_0106300 |
| chr1 | 267,883 | atp6 | E431 | Glu431Lys | atp6-E431K | No | PF3D7_0106300 |
| chr1 | 266,868 | atp6 | S769 | Ser769Asn | atp6-S769N | No | PF3D7_0106300 |
| chr7 | 403,621 | crt | N75 | Asn75Glu | crt-N75E | No | PF3D7_0709000 |
| chr7 | 404,836 | crt | Q271 | Gln271Glu | crt-Q271E | No | PF3D7_0709000 |
| chr7 | 403,700 | crt | C101 | Cys101Phe | crt-C101F | No | PF3D7_0709000 |
| chr7 | 403,620 | crt | M74 | Met74Ile | crt-M74I | No | PF3D7_0709000 |
| chr7 | 405,838 | crt | R371 | Arg371Ile | crt-R371I | No | PF3D7_0709000 |
| chr7 | 404,010 | crt | F145 | Phe145Ile | crt-F145I | No | PF3D7_0709000 |
| chr7 | 403,688 | crt | H97 | His97Leu | crt-H97L | No | PF3D7_0709000 |
| chr7 | 404,407 | crt | A220 | Ala220Ser | crt-A220S | No | PF3D7_0709000 |
| chr7 | 403,612 | crt | C72 | Cys72Ser | crt-C72S | No | PF3D7_0709000 |
| chr7 | 405,362 | crt | N326 | Asn326Ser | crt-N326S | No | PF3D7_0709000 |
| chr7 | 405,600 | crt | I356 | Ile356Thr | crt-I356T | No | PF3D7_0709000 |
| chr7 | 403,687 | crt | H97 | His97Tyr | crt-H97Y | No | PF3D7_0709000 |

| | | | | | | | |
|---|---|---|---|---|---|---|---|
| chr7 | 403,625 | crt | K76 | Lys76Thr | crt-K76T | No | PF3D7_0709000 |
| chr7 | 403,675 | crt | T93 | Thr93Ser | crt-T93S | No | PF3D7_0709000 |
| chr7 | 404,401 | crt | I218 | Ile218Phe | crt-I218F | No | PF3D7_0709000 |
| chr7 | 405,591 | crt | G353 | Gly353Val | crt-G353V | No | PF3D7_0709000 |
| chr7 | 405,560 | crt | M343 | Met343Leu | crt-M343L | No | PF3D7_0709000 |
| chrM | 4,294 | cytb | Y268 | Tyr268Cys | cytb-Y268C | No | mal_mito_3 |
| chrM | 3,890 | cytb | M133 | Met133Ile | cytb-M133I | No | mal_mito_3 |
| chrM | 4,341 | cytb | V284 | Val284Lys | cytb-V284K | No | mal_mito_3 |
| chrM | 4,293 | cytb | Y268 | Tyr268Asn | cytb-Y268N | No | mal_mito_3 |
| chrM | 4,294 | cytb | Y268 | Tyr268Ser | cytb-Y268S | No | mal_mito_3 |
| chr4 | 748,239 | dhfr-ts | N51 | Asn51Ile | dhfr-ts-N51I | No | PF3D7_0417200 |
| chr4 | 748,577 | dhfr-ts | I164 | Ile164Leu | dhfr-ts-I164L | No | PF3D7_0417200 |
| chr4 | 748,410 | dhfr-ts | S108 | Ser108Asn | dhfr-ts-S108N | No | PF3D7_0417200 |
| chr4 | 748,262 | dhfr-ts | C59 | Cys59Arg | dhfr-ts-C59R | No | PF3D7_0417200 |
| chr4 | 748,410 | dhfr-ts | S108 | Ser108Thr | dhfr-ts-S108T | No | PF3D7_0417200 |
| chr4 | 748,134 | dhfr-ts | A16 | Ala16Val | dhfr-ts-A16V | No | PF3D7_0417200 |
| chr8 | 549,685 | dhps | G437 | Gly437Ala | dhps-A437G | Yes | PF3D7_0810800 |

| | | | | | | | |
|---|---|---|---|---|---|---|---|
| chr8 | 549,681 | dhps | S436 | Ser436Ala | dhps-S436A | No | PF3D7_0810800 |
| chr8 | 549,993 | dhps | K540 | Lys540Glu | dhps-K540E | No | PF3D7_0810800 |
| chr8 | 549,682 | dhps | S436 | Ser436Phe | dhps-S436F | No | PF3D7_0810800 |
| chr8 | 549,666 | dhps | I431 | Ile431Val | dhps-I431V | No | PF3D7_0810800 |
| chr8 | 550,117 | dhps | A581 | Ala581Gly | dhps-A581G | No | PF3D7_0810800 |
| chr8 | 550,212 | dhps | A613 | Ala613Ser | dhps-A613S | No | PF3D7_0810800 |
| chr8 | 550,212 | dhps | A613 | Ala613Thr | dhps-A613T | No | PF3D7_0810800 |
| chr13 | 2,504,560 | exo | E415 | Glu415Gly | exo-E415G | No | PF3D7_1362500 |
| chr13 | 748,395 | fd | D193 | Asp193Tyr | fd-D193Y | No | PF3D7_1318100 |
| chr13 | 1,725,521 | k13 | Y493 | Tyr493His | k13-Y493H | No | PF3D7_1343700 |
| chr13 | 1,725,570 | k13 | M476 | Met476Ile | k13-M476I | No | PF3D7_1343700 |
| chr13 | 1,725,370 | k13 | I543 | Ile543Thr | k13-I543T | No | PF3D7_1343700 |
| chr13 | 1,725,382 | k13 | R539 | Arg539Thr | k13-R539T | No | PF3D7_1343700 |
| chr13 | 1,725,316 | k13 | R561 | Arg561His | k13-R561H | No | PF3D7_1343700 |
| chr13 | 1,725,259 | k13 | C580 | Cys580Tyr | k13-C580Y | No | PF3D7_1343700 |
| chr13 | 1,724,974 | k13 | A675 | Ala675Val | k13-A675V | No | PF3D7_1343700 |

| | | | | | | | |
|---|---|---|---|---|---|---|---|
| chr10 | 950,343 | kelch10 | P623 | Pro623Thr | kelch10-P623T | No | PF3D7_1022600 |
| chr13 | 2,728,402 | mcp | N252 | Asn252Asp | mcp-N252D | No | PF3D7_1368700 |
| chr5 | 960,989 | mdr1 | S1034 | Ser1034Cys | mdr1-S1034C | No | PF3D7_0523000 |
| chr5 | 961,013 | mdr1 | N1042 | Asn1042Asp | mdr1-N1042D | No | PF3D7_0523000 |
| chr5 | 958,440 | mdr1 | Y184 | Tyr184Phe | mdr1-Y184F | No | PF3D7_0523000 |
| chr5 | 961,625 | mdr1 | D1246 | Asp1246Tyr | mdr1-D1246Y | No | PF3D7_0523000 |
| chr5 | 958,145 | mdr1 | N86 | Asn86Tyr | mdr1-N86Y | No | PF3D7_0523000 |
| chr14 | 1,956,225 | mdr2 | T484 | Thr484Ile | mdr2-T484I | No | PF3D7_1447900 |
| chr13 | 958,469 | PF3D7-1322700 | T236 | Thr236Ile | PF3D7-1322700-T236I | No | PF3D7_1322700 |
| chr14 | 2,098,642 | PF3D7-1451200 | N71 | Asn71Asn | PF3D7-1451200-N71N | No | PF3D7_1451200 |
| chr4 | 881,071 | PI4K | S915 | Ser915Gly | PI4K-S915G | No | PF3D7_0419900 |
| chr7 | 896,660 | pib7 | C1484 | Cys1484Phe | pib7-C1484F | No | PF3D7_0720700 |
| chr10 | 490,720 | pph | V1157 | Val1157Leu | pph-V1157L | No | PF3D7_1012700 |
| chr6 | 1,066,989 | Sec14 | N615 | Asn615Asp | Sec14-N615D | No | PF3D7_0626400 |

**Table S3. Participant demographics of sequenced individuals in Zanzibar and mainland Tanzania.** Fourteen participants of the 5,202 genotyped did not have a case report form returned with their packet, resulting in 5,186 available for summation.

| | Total (N=5188) | Mainland (N=3755) | Zanzibar (N=1433) |
|---|---|---|---|
| **Gender** | | | |
| Female | 2471 (47.7%) | 1999 (53.3%) | 472 (33.0%) |
| Male | 2710 (52.3%) | 1750 (46.7%) | 960 (67.0%) |
| Missing | 7 | 6 | 1 |
| **Age** | | | |
| Under 5 | 1038 (20.0%) | 926 (24.7%) | 112 (7.8%) |
| 5 to 14 | 965 (18.6%) | 775 (20.6%) | 190 (13.3%) |
| 15 to 24 | 1472 (28.4%) | 986 (26.3%) | 486 (33.9%) |
| 25 to 39 | 1060 (20.4%) | 641 (17.1%) | 419 (29.2%) |
| 40 and older | 653 (12.6%) | 427 (11.4%) | 226 (15.8%) |
| **Occupation** | | | |
| Child (no occupation) | 2213 (43.1%) | 1872 (50.3%) | 341 (24.0%) |
| Farming | 951 (18.5%) | 803 (21.6%) | 148 (10.4%) |
| Trader/business | 470 (9.1%) | 186 (5.0%) | 284 (20.0%) |
| Housewife | 371 (7.2%) | 246 (6.6%) | 125 (8.8%) |
| Other | 357 (6.9%) | 178 (4.8%) | 179 (12.6%) |
| Student | 291 (5.7%) | 224 (6.0%) | 67 (4.7%) |
| Fishing | 144 (2.8%) | 73 (2.0%) | 71 (5.0%) |
| Food service | 114 (2.2%) | 78 (2.1%) | 36 (2.5%) |
| Watchman/security | 92 (1.8%) | 21 (0.6%) | 71 (5.0%) |
| Public servant/NGO | 72 (1.4%) | 30 (0.8%) | 42 (3.0%) |
| Tourism | 45 (0.9%) | 10 (0.3%) | 35 (2.5%) |
| Construction | 20 (0.4%) | 0 (0%) | 20 (1.4%) |
| Missing | 48 | 34 | 14 |
| **Parasite Density (p/µL)** | | | |
| Median (IQR) | 5,540 (1,490, 17,800) | 6,660 (1,710, 21,000) | 3,810 (1,060, 11,700) |
| Missing | 21 | 18 | 3 |

**Table S4. K13 polymorphisms detected in Zanzibar and Mainland Tanzania**

| Mutation | ZB-Travelers | ZB-NonTravelers | mTZ |
|---|---|---|---|
| **K13-P441L** | **0.002 (1/411)**<br>95% CI: 0-0.013 | 0 (0/643)<br>95% CI: 0-0.006 | **0.001 (3/2855)**<br>95% CI: 0-0.003 |
| **K13-R561H** | 0 (0/364)<br>95% CI: 0-0.01 | 0 (0/585)<br>95% CI: 0-0.006 | **0.001 (3/2598)**<br>95% CI: 0-0.003 |
| **K13-A675V** | 0 (0/470)<br>95% CI: 0-0.008 | **0.001 (1/729)**<br>95% CI: 0-0.008 | 0 (0/3215)<br>95% CI: 0-0.001 |

**Table S5. Prevalence of biallelic single nucleotide polymorphisms in antimalarial resistance genes determined by molecular inversion probes in districts in mainland Tanzania and Zanzibar**

See uploaded excel document.

**Table S6. Publicly Available Genomes Used in Analysis**

| P441L | R561H | A675V | Wild Type |
|---|---|---|---|
| PD0478-C | PD0068-C | PD0462-C | ERR4283089 |
| PD0491-C | PD0079-C | PD0471-C | ERR4283090 |
| PD0552-C | PD0126-C | PD0480-C | ERR4283091 |
| PD0556-C | PD0481-C | PD0492-C | ERR4283092 |
| PD0563-C | PD0495-C | PD0497-C | ERR4283093 |
| PD0564-C | PD0562-C | PD0512-C | ERR4283094 |
| PD0566-C | PD0565-C | PD0513-C | ERR4283095 |
| PD0581-C | PD0799-C | PD0522-C | ERR4283096 |
| PD0798-C | PD0800-C | PD0529-C | ERR4283097 |
| PD0808-C | PD0819-C | PD0548-C | ERR4283098 |
| PD0809-C | PD0822-C | PD0557-C | ERR4283099 |
| PD0810-C | PD0823-C | PD0627-C | ERR4283100 |
| PD0820-C | PD0824-C | PD0746-C | ERR4283101 |
| PD0830-C | PD0826-C | PD0867-C | ERR4283102 |
| PD0870-C | PD0828-C | PD0898-C | ERR4283103 |
| PD0887-C | PD0832-C | PD0924-C | ERR4283104 |
| PD1003-C | PD0876-C | PD0945-C | ERR4283105 |
| PD1091-C | PD0877-C | PD0989-C | ERR4283106 |
| PD1095-C | QC0184-C | PD1001-C | ERR4283107 |
| PD1096-C | QC0185-C | PD1059-C | ERR4283108 |
| PD1155-C | QC0250-C | PD1098-C | ERR4283109 |
| QC0133-C | RCN03272 | PD1126-C | ERR4283110 |
| QC0152-C | RCN03319 | PD1185-C | ERR4283111 |
| QC0154-C | RCN03339 | PD1193-C | ERR4283112 |
| QC0165-C | RCN03348 | PD1196-C | ERR4283113 |
| QC0186-C | RCN03388 | PD1197-C | MSMTA1 |
| QC0209-C | RCN03502 | PD1237-C | MSMTA10 |
| QC0214-C | RCN03510 | QC0129-C | MSMTA11 |
| QC0223-C | RCN03535 | QC0160-C | MSMTA12 |
| RCN03306 | RCN09301 | DRA005348 | MSMTA2 |
| RCN03317 | RCN09302 | ERS7266441 | MSMTA3 |
| RCN03326 | RCN09312 | | MSMTA4 |
| RCN03343 | RCN09313 | | MSMTA5 |
| RCN03351 | RCN09314 | | MSMTA6 |
| RCN03367 | RCN09316 | | MSMTA7 |
| RCN03376 | RCN09317 | | MSMTA8 |
| RCN03385 | RCN09319 | | MSMTA9 |
| RCN03409 | RCN09326 | | MSMTB1 |
| RCN03422 | RCN09329 | | MSMTB10 |
| RCN03423 | RCN09340 | | MSMTB11 |
| RCN03425 | RCN09399 | | MSMTB12 |
| RCN03426 | RCN09402 | | MSMTB2 |
| RCN03474 | RCN09466 | | MSMTB3 |
| RCN03485 | RCN09486 | | MSMTB4 |
| RCN03489 | RCN09489 | | MSMTB5 |
| RCN03490 | RCN09503 | | MSMTB8 |
| RCN03496 | RCN09544 | | MSMTB9 |
| RCN03504 | RCN09571 | | MSMTC1 |
| RCN03511 | RCN09580 | | MSMTC10 |
| RCN03515 | RCN09592 | | MSMTC11 |
| RCN03536 | RCN09602 | | MSMTC12 |
| RCN09334 | RCN09658 | | MSMTC2 |
| RCN09367 | RCN09677 | | MSMTC4 |

| | | | |
|---|---|---|---|
| RCN09370 | RCN09694 | | MSMTC5 |
| RCN09418 | RCN09716 | | MSMTC8 |
| RCN09419 | RCN09812 | | MSMTC9 |
| RCN09439 | RCN09828 | | MSMTD1 |
| RCN09472 | RCN09832 | | MSMTD10 |
| RCN09493 | RCN09845 | | MSMTD11 |
| RCN09507 | RCN09935 | | MSMTD12 |
| RCN09525 | RCN09954 | | MSMTD2 |
| RCN09527 | RCN09963 | | MSMTD3 |
| RCN09540 | RCN09964 | | MSMTD4 |
| RCN09542 | RCN09979 | | MSMTD5 |
| RCN09550 | RCN09989 | | MSMTD6 |
| RCN09554 | RCN10002 | | MSMTD7 |
| RCN09567 | RCN10007 | | MSMTD8 |
| RCN09572 | RCN10008 | | MSMTD9 |
| RCN09574 | RCN10009 | | MSMTE12 |
| RCN09577 | RCN10019 | | MSMTF4 |
| RCN09582 | RCN12401 | | MSMTG6 |
| RCN09584 | RCN12402 | | MSMTH1 |
| RCN09587 | RCN12403 | | MSMTH10 |
| RCN09589 | RCN12404 | | MSMTH12 |
| RCN09590 | PRJEB38946 | | MSMTH2 |
| RCN09607 | PRJNA1092065 | | MSMTH3 |
| RCN09611 | PRJNA119461 | | MSMTH4 |
| RCN09619 | | | MSMTH5 |
| RCN09625 | | | MSMTH6 |
| RCN09630 | | | MSMTH7 |
| RCN09636 | | | MSMTH8 |
| RCN09638 | | | PD0009-01 |
| RCN09655 | | | PD0031-C |
| RCN09656 | | | PD0032-C |
| RCN09657 | | | PD0034-C |
| RCN09687 | | | PD0038-C |
| RCN09703 | | | PD0040-C |
| RCN09709 | | | PD0041-C |
| RCN09729 | | | PD0042-C |
| RCN09735 | | | PD0043-C |
| RCN09736 | | | PD0046-C |
| RCN09740 | | | PD0048-C |
| RCN09744 | | | PD0050-C |
| RCN09756 | | | PD0052-C |
| RCN09780 | | | PD0053-C |
| RCN09783 | | | PD0056-C |
| RCN09792 | | | PD0057-C |
| RCN09818 | | | PD0058-C |
| RCN09835 | | | PD0061-C |
| RCN09837 | | | PD0062-C |
| RCN09852 | | | PD0070-C |
| RCN09855 | | | PD0072-C |
| RCN09857 | | | PD0073-C |
| RCN09868 | | | PD0074-C |
| RCN09881 | | | T0901AD0 |
| RCN09913 | | | T0901BD0 |
| RCN09915 | | | T0902AD0 |
| RCN09916 | | | T0902BD0 |

| | | | |
|---|---|---|---|
| RCN09921 | | | T0903AD0 |
| RCN09924 | | | T0903BD0 |
| RCN09926 | | | T0904AD0 |
| RCN09927 | | | T0904BD0 |
| RCN09936 | | | T0905AD0 |
| RCN09939 | | | T0905BD0 |
| RCN09943 | | | T0906AD0 |
| RCN09951 | | | T0906AD21 |
| RCN09953 | | | T0906BD0 |
| RCN09959 | | | T0907AD0 |
| RCN09960 | | | T0907BD0 |
| RCN09961 | | | T0908AD0R |
| RCN09962 | | | T0908BD0R |
| RCN09965 | | | T0909AD0 |
| RCN09967 | | | T0909BD0 |
| RCN09970 | | | T0910AD0 |
| RCN09974 | | | T0910BD0 |
| RCN09978 | | | T0911AD0 |
| RCN10006 | | | T0911BD0 |
| | | | T0912AD0 |
| | | | T0912BD0 |
| | | | T0913AD0 |
| | | | T0913BD0 |
| | | | T0914AD0 |
| | | | T0915AD0 |
| | | | T0915BD0 |
| | | | T0916AD0 |
| | | | T0916BD0 |
| | | | T0917AD0 |
| | | | T0917BD0 |
| | | | T0918AD0 |
| | | | T0918BD0 |
| | | | T0919AD0 |
| | | | T0919BD0 |
| | | | T0920AD0 |
| | | | T0920BD0 |
| | | | T0921AD0 |
| | | | T0921BD0 |
| | | | T0922AD0 |
| | | | T0922BD0 |
| | | | T0923AD0 |
| | | | T0923BD0 |
| | | | T0924AD0 |
| | | | T0925AD0 |
| | | | T0925BD0 |
| | | | T0926AD0 |
| | | | T0926BD0 |
| | | | T0927AD0 |
| | | | T0927BD0 |
| | | | T0928AD0 |
| | | | T0929AD0 |
| | | | T0930AD0 |
| | | | T0930BD0 |
| | | | T0931AD0 |
| | | | T0932AD0 |

| | | | |
|---|---|---|---|
| | | | T0933AD0 |
| | | | T0936AD0 |
| | | | T0936AD28 |
| | | | T0937BD0 |
| | | | T0938AD0 |
| | | | T0939BD0 |
| | | | T0940AD0 |
| | | | T0943BD0 |
| | | | T0944AD0 |
| | | | T0945AD0 |
| | | | T0946BD0 |
| | | | T0949BD0 |
| | | | T0952AD14 |
| | | | T0955BD0 |
| | | | T0956BD0 |
| | | | T0957BD0 |
| | | | T0961AD0 |
| | | | T0962AD0 |
| | | | T0964AD0 |
| | | | T0967AD0 |
| | | | T0967BD0 |
| | | | T0968BD0 |
| | | | T0973AD0 |
| | | | T0973BD0 |
| | | | T0980AD0 |
| | | | T0982AD0 |
| | | | T0982AD21 |
| | | | T0983BD0 |
| | | | T0985AD0 |
| | | | T0987AD0 |
